# A delayed SEIQR epidemic model of COVID-19 in Tokyo area

**DOI:** 10.1101/2020.08.18.20177709

**Authors:** Kazuo Maki

**Affiliations:** MediEco R & D Corporation, Hieidaira, Ohtsu 520-0016, Japan

## Abstract

The infection of COVID-19 has caused a global pandemic. In order to avoid excessive restriction to the social activity, a good strategy of quarantine based on a realistic model is expected. Several epidemic models with a quarantine compartment such as susceptible-exposed-infectious-quarantined-recovered (SEIQR) model have been applied. However, in the actual situation, the infection test and quarantine is often delayed from the beginning of the infectious stage.

This article presents a delayed SEIQR model to analyze the effect of the delay of quarantine. The latency period (compartment E) and the incubation period were assumed to be 3 days and 5 days, respectively. Considering that the presymptomatic infection ratio is 0.4, the natural decay rate of the number of infectious patients was assumed to be 0.25 days^-1^. The recovery rate was assumed to be 0.07 days^-1^ from the typical PCR test positive period. The PCR test positive number in the period from March 10 to July 18 in 2020 in Tokyo area was analyzed. The delay time distribution of the quarantine was derived from the record of the symptom onset date, and was utilized to determine the delay time profile of quarantine in the model.

It was found that the major contributor to the infection control was the restraint of social contact. However, the quarantine action also contributed to reducing the reproduction number by the ratio of 0.88 and 0.8 in the period from March 10 to June 3 and from June 4 to July 18, respectively. The delay of quarantine was found to be well correlated to the effectiveness of quarantine. Therefore, a record on a symptom onset date is very important to estimate the effect of quarantine measure. The basic reproduction number was estimated to be 2.56. In view of the presymptomatic infection ratio 0.4, it would be very hard to restrain the expansion of infection only by quarantining the symptomatic patients.

## 1. Introduction

The novel coronavirus disease (COVID-19) has caused the infection expansion all over the world in the second quarter of 2020. A deterministic mathematical modeling of the outbreak of infection is expected to be a useful tool to analyze the situation and to suggest a plausible measure. In the SIR model [1], the total population *N* of the area under consideration is divided into three compartments: susceptible (*S*), infectious (*I*), and recovered (*R*) individuals. In the SEIR model [2], another compartment, exposed individuals (*E*), is added to take a latency period into account. However, COVID-19 is different from most conventional infection cases in the point that unique symptom is not well established yet and that asymptomatic patients may be infectious [3],[4]. Since an infectious patient cannot be identified clearly, we have to restrain contact between the others in daily life. Influence to social activities is enormous. It is necessary to find and quarantine an infectious patient by some kind of test, and this quarantine measure deeply affects the situation of the infection expansion.

Models with a compartment for quarantined individuals are called SIQR model [5] or SEIQR model [6],[7],[8]. The occurrence of the transition from the component I to Q, which is observed as a number of newly found infected patients, is usually assumed to be proportional to the number of infectious patients at large, and this proportionality coefficient is treated as a parameter called “quarantine rate”. The outbreak of COVID-19 has been analyzed by many authors in terms of quarantine [9],[10],[11],[12],[13],[14]. The quarantine rate was estimated from available data under some simple assumptions or using statistical method [10] or AI model [11]. However, infected patients usually have a PCR examination after developing a symptom. Moreover when a check system is not well coordinated, they have to stand by for several days. Young et al. developed a delayed SEIQR model [15] including this delay effect, which was applied to COVID-19 by Vysarayani and Chatterjee [16]. In their model, the patient that has passed an assumed period is quarantined with an assumed probability, but if not quarantined, a chance of being quarantined is not left any longer. On the other hand, Utamura et al. [17] developed a model in that all the infectious patients are quarantined after passing an assumed period (14 days).

In this report, I will present a new model with two compartments of infectious patients: a quarantine waiting compartment and a quarantine possible compartment. Patients are quarantined with a quarantine rate only in the latter compartment. The quarantine rate defined in this model is a natural extension of that of non-delayed SIQR or SEIQR models.

In the next section, a delayed SEIQR model will be formulated. In Section 3, the data of the daily found PCR positive patients in Tokyo area [18] will be analyzed. The last section will be devoted to a discussion and conclusion.

## 2. A delayed SEIQR model

In this section, a delayed SEIQR model will be formulated. As a compartment model, this model is similar to the delayed SEIQR model proposed by Young et al. [15]. As Young’s model, I assumed that a patient stays in the compartment E for a definite period *ε*. As for the compartment I, which is the point of difference to Young’s model, this compartment was divided into J and K, and only those belonging to K were assumed to be quarantined. The patient stays in the compartment J for a period *τ* and automatically moves to the compartment K. Here, a quarantine rate *q* was introduced as an extension of that of non-delayed SIQR or SEIQR model. All the members in K were assumed to be continuously quarantined with a rate *q* if not recovered. The parameter *τ* means a delay of the start of quarantine. If we expand the test to check earlier stage patient, *τ* can be decreased. In the case of non-delayed models (τ=0), a constant value is imposed on *q* for the whole stage of an infectious patient at large. The flow chart is shown in Fig.1.

**Figure 1:**
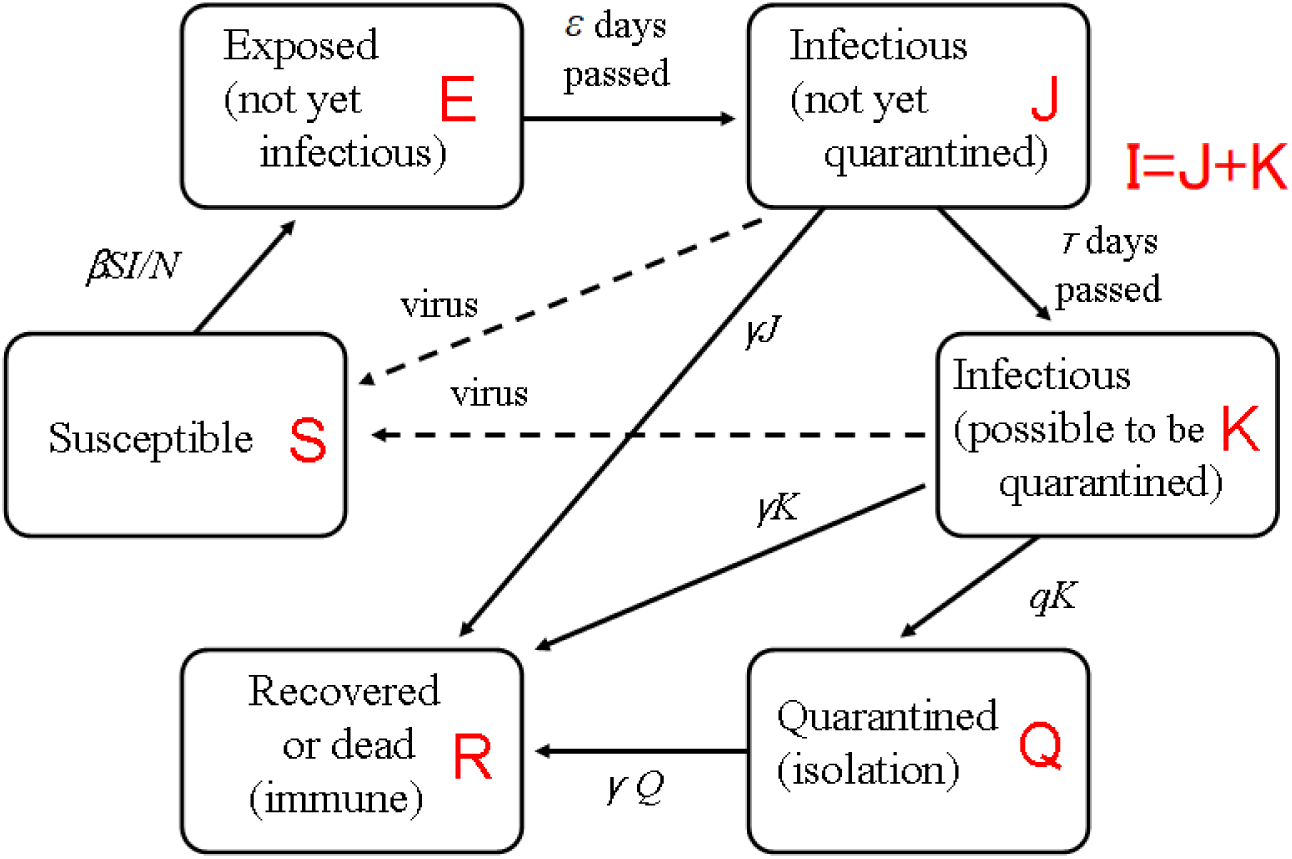
Flow chart of a delayed SEIQR model.

Thus, the delay differential equations are given as follows. (The time dependence of coefficients *β* and *q* is not indicated for simplicity.)

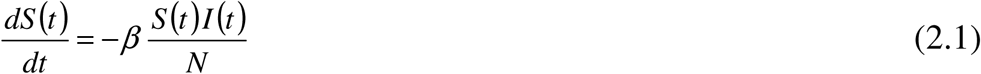

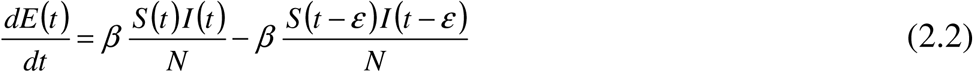

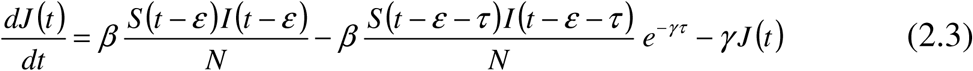

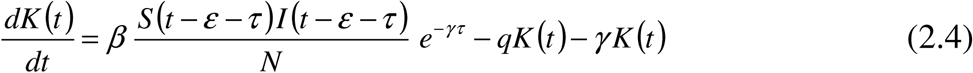

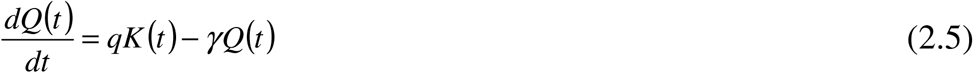

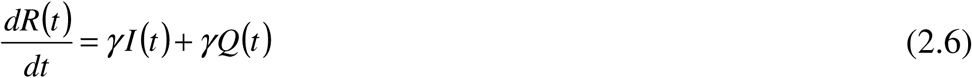

where

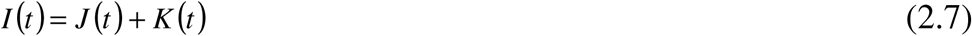

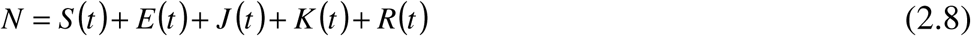

The parameters *β, γ, q, ε*, and *τ* represent the transmission rate, the natural decay rate of the number of infectious patients, the quarantine rate, the latency period, and the delay of the start of quarantine, respectively. The transmission rate may be interpreted as the frequency of daily contact with others multiplied by the transmission risk from an encountered infectious patient. Birth and death processes are neglected here. The factor exp(-*γτ*) in Eqs. (2.3) and (2.4) is necessary to take into account the decay with the rate *γ* in the compartment J.

It should be noted here that the PCR test positive result does not always mean infectiousness because PCR test does not detect active virus but detects only RNA [19],[20]. The recovery rate, denoted as *γ’*, is defined here as the inverse of the averaged PCR test positive period. This rate is smaller than the natural decay rate *γ* of the number of infectious patients. Thus we have to assume similar compartments J’ and K’ for PCR test positive patients as an extension of J and K, respectively. The number of PCR test positive patients in the compartment K’, denoted as *K’*(*t*), satisfies the following additional delay differential equation.

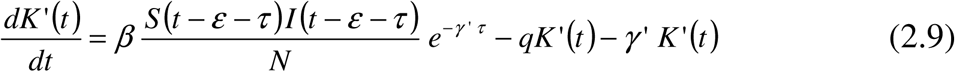

Then the number of the newly found PCR test positive patient is

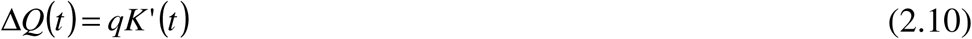

When the parameters are made *ε* =0, *τ*= 0, *ε* = *τ*= 0, *τ* =*q*=0, and *ε* = *τ* =*q*=0, this model becomes a delayed SIQR model, an SEIQR model, an SIQR model, an SEIR model, and an SIR model respectively.

Hereafter, I will consider only the early stage of outbreak. I made an approximation as *S*=*N*, and only Eqs. (2.3), (2.4), (2.7), (2.9), and (2.10) will be used. These equations are rewritten as

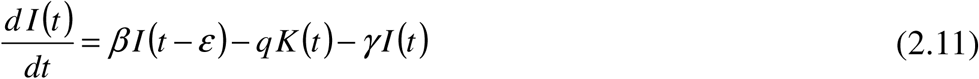

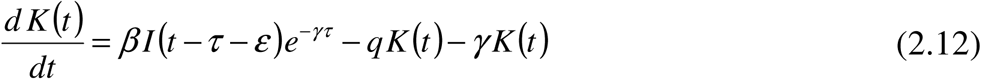

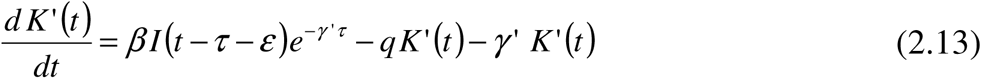

Next, I will establish the relation of the model parameters to the actual epidemic data. In addition to the PCR test positive fixation date *T*_f_, the epidemic data usually contain the symptom onset date *T*_s_ if the patient is symptomatic. The delay of quarantine is defined as *T*_d_=*T*_f_−*T*_s_+*T*_p_, where *T*_p_ is the presymptomatic infectious period. (I will assume later that *T*_p_=2 days.) In order to derive the expression of the distribution function of *T*_d_, we have to calculate the probability *P*(*t*) that a patient who became infectious at *t*=0 is not quarantined while keeping PCR test positive. Since the decay rate is *γ’* before the start of quarantine (*t*<*τ*), and then *γ’*+*q* if *t*>*τ*.

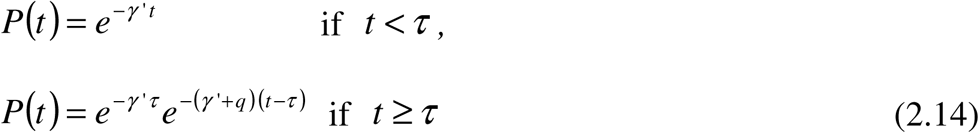

The probability of daily quarantine in the PCR test is given by *qP*(*t*) (*t* ≥ *τ*), which is rewritten as

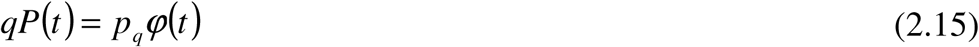

where *φ*(*t*) = 0 if *t* < *τ*,

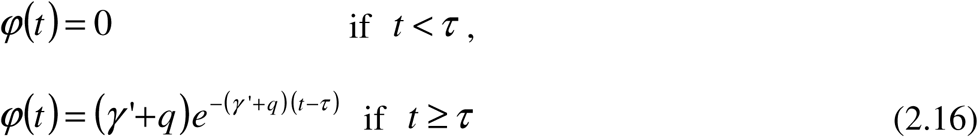

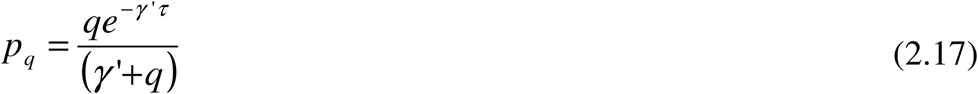

The function *φ*(*t*) and the coefficient *p*_q_ are interpreted as the normalized distribution of the delay time of quarantine and the ratio of the PCR test detection of infected patient, respectively. It should be noted that *φ*(*t*) can be observed, but *p*_q_ cannot be observed directly. Although the observed quarantine delay distribution is different from the distribution *φ*(*t*) of the model, the most important parameter is the averaged delay time <*T*_d_>. Hereafter, < > denotes an average by the observed time distribution of the delay of quarantine. Model parameters should give the same averaged delay time.

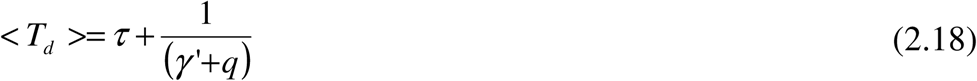

If we assume that the standard deviation *σ* should be the same as observed,

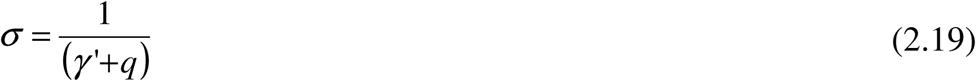

These equations determine *τ* and *q*.

If we replace *γ’* with *γ, P*(*t*) will be the probability that an infected patient is not quarantined while maintaining infectiousness. The effective reproduction number *R*_*e*_ is given by carrying out the time integration of *P*(*t*) (Eq.(2.14) where *γ’* is replaced with *γ*) multiplied by *β*.

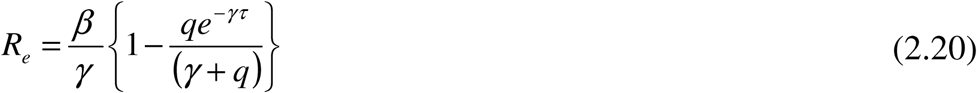

The second term in the parenthesis {} means the ratio of the infectious patient detected and quarantined by the PCR test. The ratio *R*_*e*_/(*β*/*γ*) may be interpreted as the effect of quarantine when excluding the social contact avoidance effect. This ratio is expressed in terms of the characteristic parameters of the distribution of the delay of quarantine.

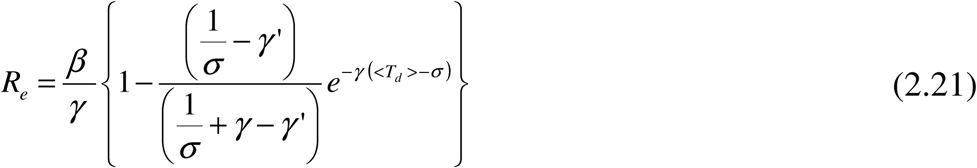

As an example, the ratio *R*_*e*_/(*β*/*γ*) is plotted in Fig.2, using the values of *γ=*0.25 days^-1^ and *γ’ =*0.25 days^-1^ as will be determined in the next section. It is well correlated with the averaged delay of quarantine. In Appendix A, *R*_*e*_ is expressed directly by an observed time distribution of the delay of quarantine in a different epidemic model.

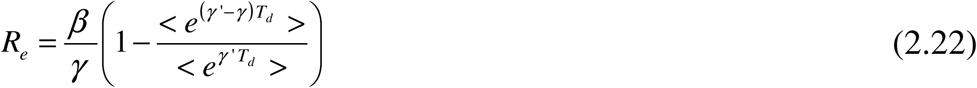

**Figure 2.**
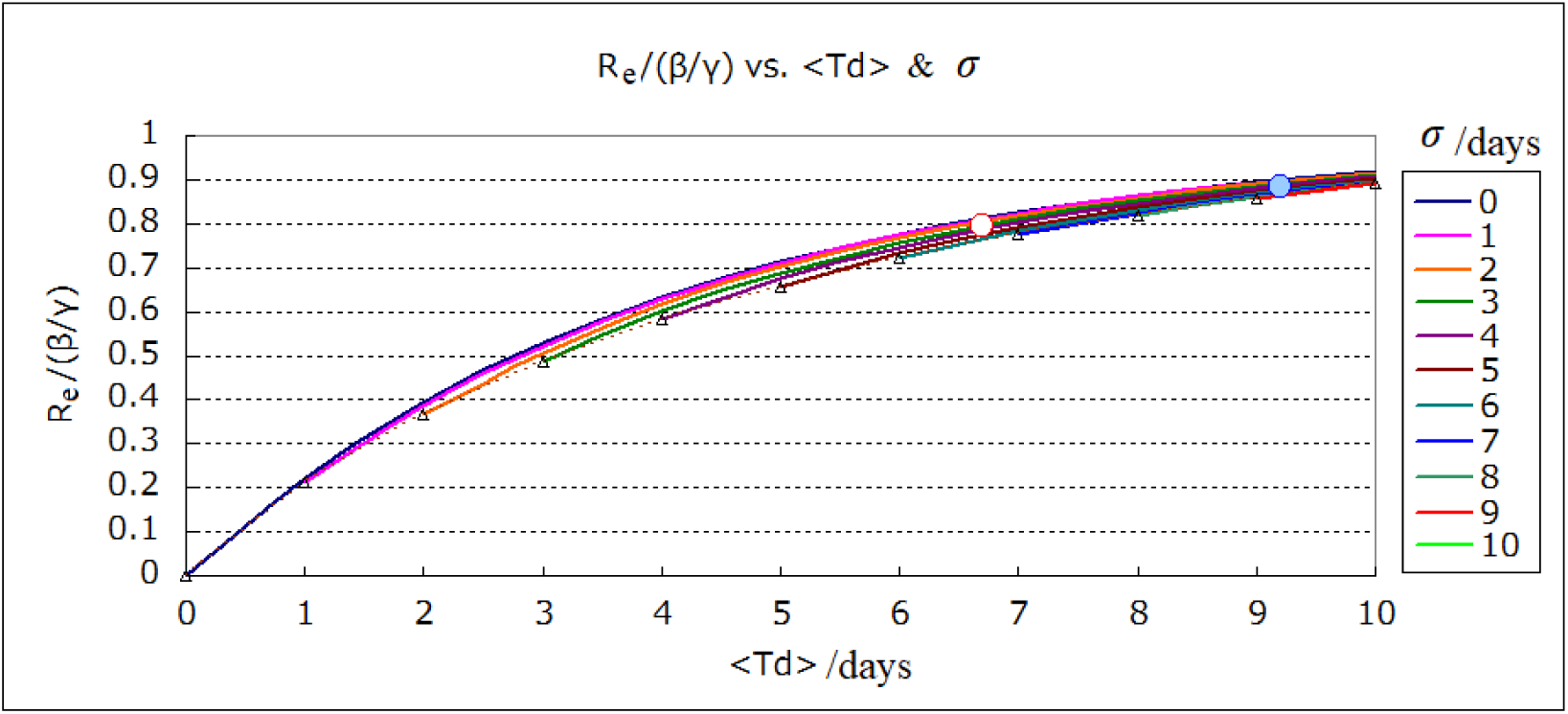
Effect of quarantine on the reproduction number. <*T*_d_> and *σ* denote an average and a standard deviation of the delay of quarantine, respectively. The small triangles correspond to the case that <*T*_d_> = *σ* or τ=0. Two circles correspond to the Tokyo area data in Sec.3.

The accuracy of the approximation in Eq. (2.21) can be evaluated by this equation.

In the case of non-delayed models (*τ*=0) the reproduction number is expressed as *R*_*e*_=*β*/(*γ+q*). This case is indicated by small triangles in Fig.2. The basic reproduction number is estimated as *R*_0_=*β*/*γ* using the earliest value in *β*.

There is a stationary solution proportional to exp(*λt*) in Eqs. (2.11) and (2.12). The coefficient *λ* is called the infection expansion coefficient, and it satisfies the following relation, which will be useful not only in finding *β* from the observed time profile of infection but also in predicting the expansion rate *λ*.

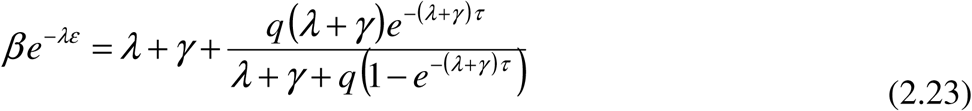

The left hand side may be interpreted as an effective transmission rate, which depends on the latency period *ε*. If the infection is expanding (*λ*>0) or shrinking (*λ*<0), the effective transmission rate is reduced or enhanced, respectively.

## 3. Analysis of the time profile of infection in Tokyo area

In this section, I will analyze the data of the number of PCR test positive person who were living and tested in Tokyo area (Tokyo, Kanagawa, Saitama, and Chiba) [18]. The test period is from March 10 to July 18 in 2020. The total number of patients is 13,978. The parameters *γ, γ’*, and *ε* will be estimated from references, and the parameters *τ* and *q* will be derived from the observed quarantine delay distribution. Finally, the parameter *β* will be chosen so as to reproduce the time profile of the number of PCR test positive patients.

The incubation period has a wide distribution and the average is about 5 days [21]. It is also known that patients are infectious 2 days before the onset of symptom [3],[4]. Thus, I assumed that the latency period *ε* is 3 days. I assumed that the natural decay rate *γ* of the number of infectious patients is 0.25 days^-1^ so as to make the ratio of presymptomatic infection 0.4 as observed [3],[4]. Since the PCR test positive period has a wide distribution [22],[23], I assumed 14 days as a typical period. The recovery rate *γ’* was set to be 1/14=0.07 days^-1^. The time profile of probability of being infectious is compared with that of being PCR test positive in Fig.3.

**Figure 3.**
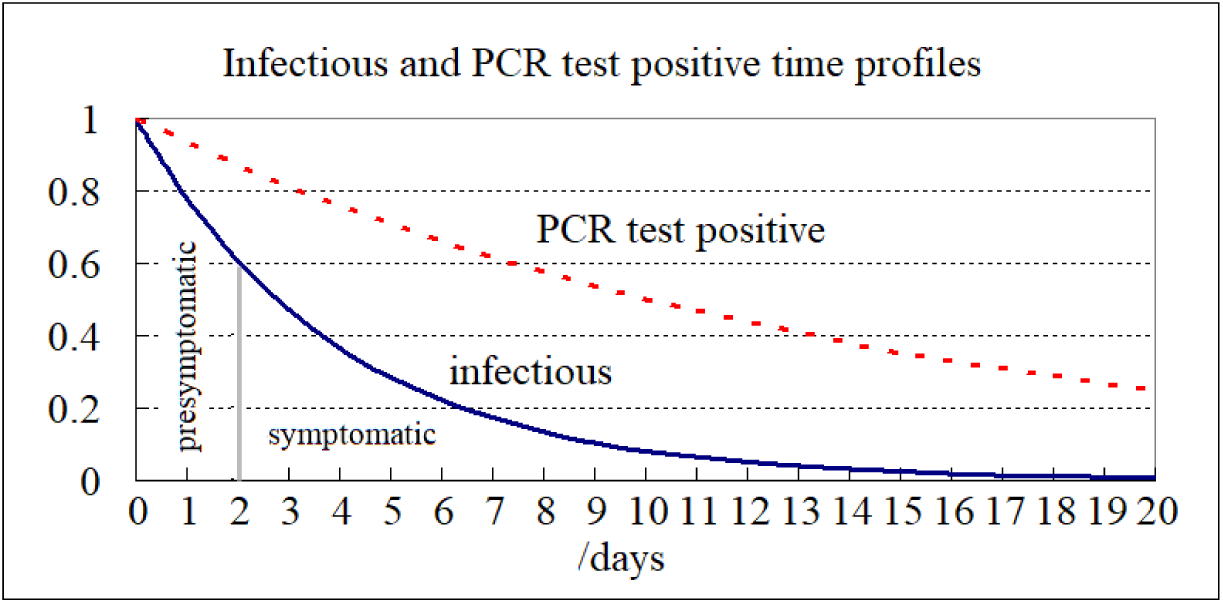
Comparison of infectious and PCR test positive time profiles.

The symptom onset date (*T*_s_) was recorded in 24% of the data of Tokyo area [18]. The daily average of the delay of quarantine (*T*_d_=*T*_f_−*T*_s_+*T*_p_) is shown in Fig.4. Although the data are scattered, the delay in June and July is apparently shorter than that in March, April, and May. I divided the whole period at the point between June 3 and 4.

**Figure 4.**
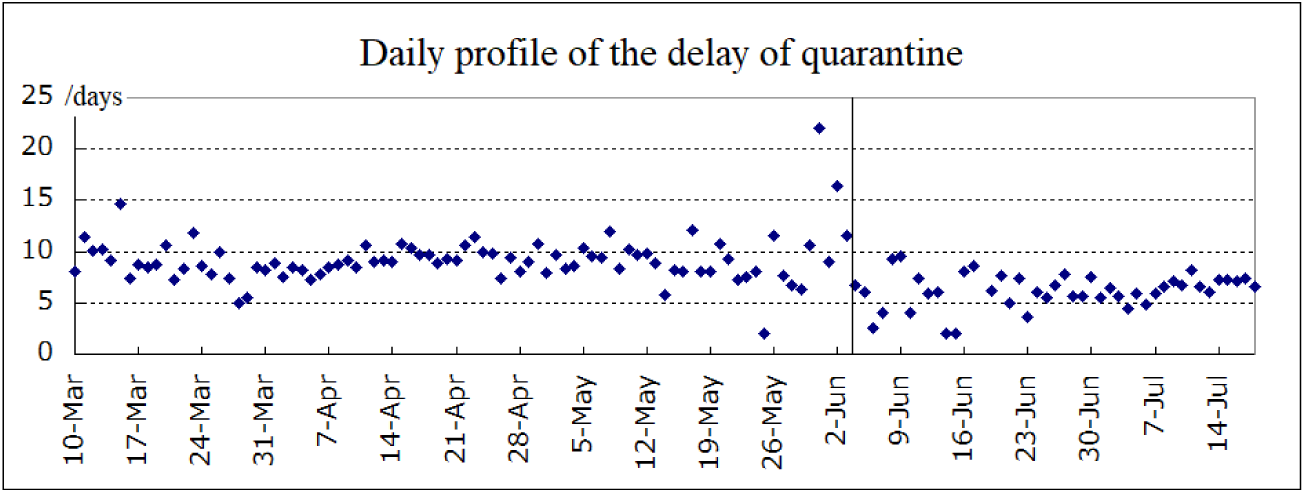
Daily profile of the delay of quarantine. The delay of the quarantine is defined as the period from the start of infectiousness to the PCR test positive fixation day. It is assumed here that the start of infectiousness is 2 days earlier than the onset of symptom.

The distributions of the delay of quarantine in these periods are shown in Fig.5. The average (<*T*_d_>) and the standard deviation (*σ*) of the delay is 9.17 days and 4.24 days respectively in the former period, and 6.69 days and 3.08 days respectively in the latter period. The parameters *τ, q*, and the effect of quarantine (*R*_*e*_/(*β*/*γ*)) were calculated by Eqs. (2.18), (2.19), and (2.20). The result was that *τ*=4.93 days, *q*=0.166 days^-1^, and *R*_*e*_/(*β*/*γ*)=0.884 in the former period, while *τ*=3.6 days and *q*=0.25 days^-1^, and *R*_*e*_/(*β*/*γ*)=0.795 in the latter period. These are indicated by two small circles in Fig.2. (In the case of the exact expression (Eq. (2.22)), *R*_*e*_/(*β*/*γ*)=0.879 and *R*_*e*_/(*β*/*γ*)=0.792 in the former and the latter period, respectively. This means that the present model is a good approximation.) Since a small change of parameters does not affect the result much as far as the effect of quarantine is the same, I applied a common value of *τ*=4 days for a convenience on the numerical calculation, and *q* was readjusted so as to keep the effect of quarantine. I obtained the values as *q*=0.116 and *q*=0.314 for the former and the latter period, respectively. If we set that *τ*=0: i.e. in the case of non-delayed model, *q*=0.039 days^-1^ and *R*_*e*_/(*β*/*γ*)=0.865 in the former period, *q*=0.0795 and *R*_*e*_/(*β*/*γ*)= 0.759 in the latter period. The delay time profiles in the two cases (*τ*=4 and *τ*=0) are compared with the observed profiles in Fig.5.

**Figure 5.**
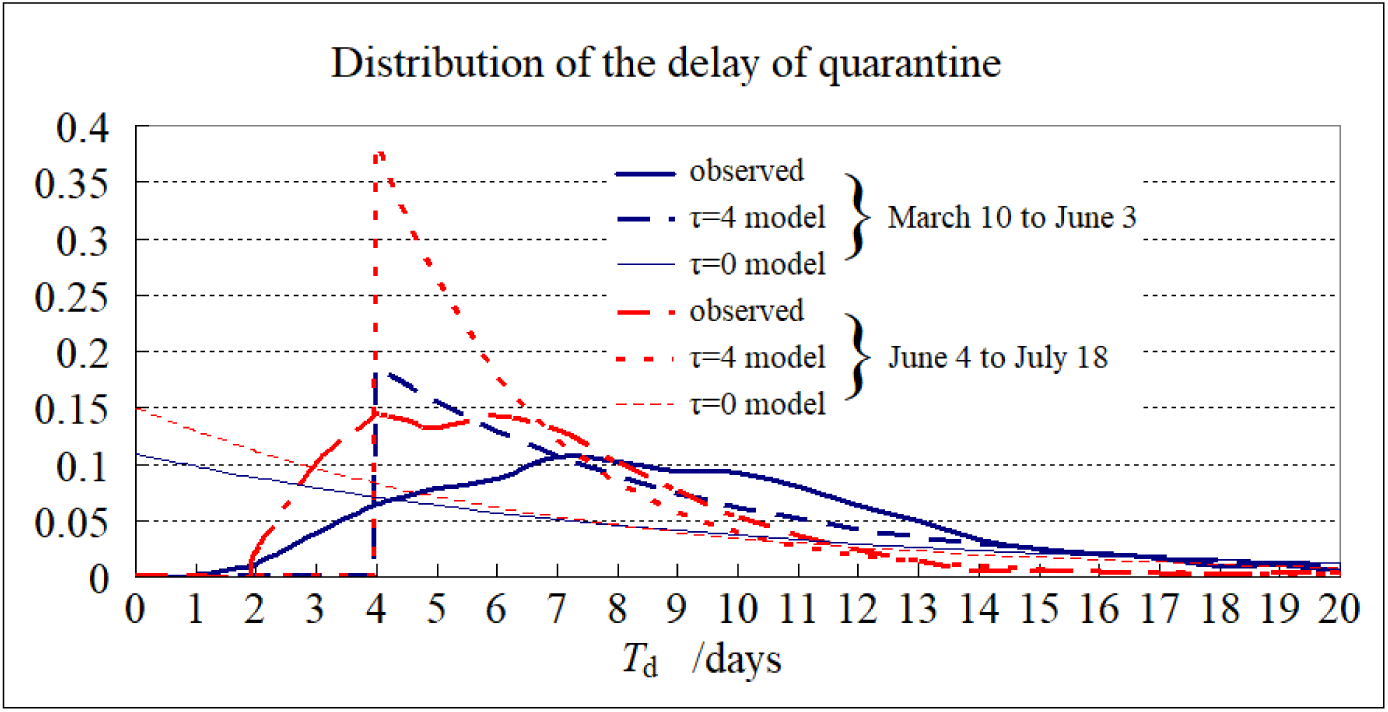
Distribution of the delay of quarantine.

Numerical calculation was made by the fourth order Runge-Kutta method with time step=0.1 days. The PCR test positive person number data of Tokyo area Δ*Q*(*t*) in the period from March 10 to July 18 [18] was reproduced by choosing the parameter *β*. The time profile of infection is shown in Fig.6. The parameters *β* and *q* and the effective reproduction number *R*_e_ in each period are listed in Table 1. As for the parameters *β* and *q*, the moving average of 7 days of the values in Table 1 was adopted as shown in Fig.6. The small protuberance in Δ*Q*(*t*) around June 3 reflects the change in the parameter *q* (the enhancement of PCR test). The basic reproduction number *R*_0_ is found to be 0.640/0.25=2.56.

**Table 1:** Parameter set and effective reproduction number. The parameter *τ* is set to be 4 days. The symbols *β, q*, and *Re* denote transmission rate (days^-1^), quarantine rate (days^-1^), and the effective reproduction number, respectively. *β*/0.640 or *Re*/(*β*/*γ*) means a contribution of *β* or that of quarantine, respectively, to the reduction ratio of the reproduction number.

| Period | a: Mar 10-Apr 1 | b: Apr 2-Apr 22 | c: Apr 23-May 15 | d: May 16-Jun 3 | e: Jun 4-Jul 18 |
| --- | --- | --- | --- | --- | --- |
| $\beta$ | 0.640 | 0.180 | 0.146 | 0.329 | 0.474 |
| $q$ | 0.116 | 0.116 | 0.116 | 0.116 | 0.314 |
| $R_e$ | 2.26 | 0.64 | 0.52 | 1.16 | 1.51 |
| $\beta/0.640$ | 1 | 0.28 | 0.23 | 0.51 | 0.74 |
| $R_e/(\beta/\gamma)$ | 0.884 | 0.884 | 0.884 | 0.884 | 0.795 |

The reduction ratio of the reproduction number is a product of the social action inhibiting effect (*β*/0.640) and the quarantine effect (*R*_*e*_/(*β*/*γ*)). They are shown in Table.1. The effect of the social behavior restraint is dominant on the whole. It is remarkable in the period b and c in particular. Death of a famous comedy actor was reported, and people checked social behavior. An emergency declaration was issued, and social contact restraint of 80% was recommended. After the emergency declaration was released, *β* increased in the period d and e, but the PCR check system has been strengthened by period e by the ratio 0.204/0.115=1.77. This eased infection expansion. The time profile of infection after those periods is not shown here. The social behavior was limited again, and infection calmed down a little throughout August and September. However, in view of the economical influence, the quarantine action should be strengthened.

In order to grasp the situation, the curves which make the infection expansion rate *λ* fixed (Eqs. (2.18), (2.19), and (2.23)) are drawn in the plane of *β* and <*T*_d_> in Fig.7. (Note that *λ*=−0.05 or −0.1 means 30% or 50% decrease of infection a week, respectively.) Since there is a small dependence on *σ*, the two extreme cases are drawn. One is the case that *q*=∞ (*σ*=0; i.e. <*T*_d_>=*τ*), the other is the case that *τ*=0 (<*T*_d_>=*σ*: i.e. non-delayed SEIQR model). The result of the analysis in Tokyo area is indicated by the grey dots. As an example, let us start from the period e. If the quarantine measure is the same, we have to restrain the social activity by 35% to get over the *λ*=0 line (*β*=0.31 day^-1^), but if the quarantine measure could be strengthened as that <*T*_d_>=4 days, only 16% reduction of *β* would be sufficient to get over the *λ*=0 line (*β*=0.4 day^-1^). The economic impact would be much mitigated. However, when only the symptomatic patient is quarantined, <*T*_d_> cannot be made shorter than 2 days. This also means that *R*_e_ cannot be made less than 1.04=0.4*R*_0_, if we recover the level of social activity before the outbreak of COVID-19. (The number 0.4 is the presymptomatic ratio of infection.) Thus the quarantine of presymptomatic patient is necessary.

## 4. Summary and discussion

Conventional SIQR or SEIQR (non-delayed) models assume that the number of quarantined patients daily is proportional to the number of infectious patients at large. As shown by thin lines in Fig.5, this supposition is equivalent to assuming an exponential distribution of the quarantine delay from the beginning of infectiousness. However, the observed delay time distribution looks very different from that assumed in these models. In order to reflect the essential characteristics of the distribution, I expanded conventional SEIQR models with a new parameter *τ* describing the delay of the start of quarantine. I also estimated the natural decay rate *γ* of the number of infectious patients from the observed ratio of presymptomatic infection. The decay rate was found to be much larger than usual estimates from medical recovery data. Then I introduced a recovery rate *γ’* as the inverse of a PCR test positive period to reproduce the observed PCR test positive number.

I found that the quarantine effect on the reduction of the effective reproduction number is closely related to the averaged delay of quarantine as is shown in Fig.2. This means that we can estimate the effect of quarantine measure directly. (The exact relation is given in Eq. (2.22).) Although this estimation from the averaged delay is also possible in the non-delayed models, the estimation itself tends to be rather optimistic. It should also be noted that, in the non-delayed models, a prediction in terms only of *β* and *q* might become too optimistic because the difficulty of shortening the delay of quarantine is often underestimated.

The infection expansion or shrinking can be predicted in Fig.7, where the infection expansion rate *λ* is expressed in terms of the transmission rate *β* and the averaged delay of quarantine <*T*_d_>. As a point of the infection measure, I share a view with He et al. [3] and Chun et al. [4]. The quarantine of presymptomatic patient is necessary to recover the normal social activities.

**Figure 6:**
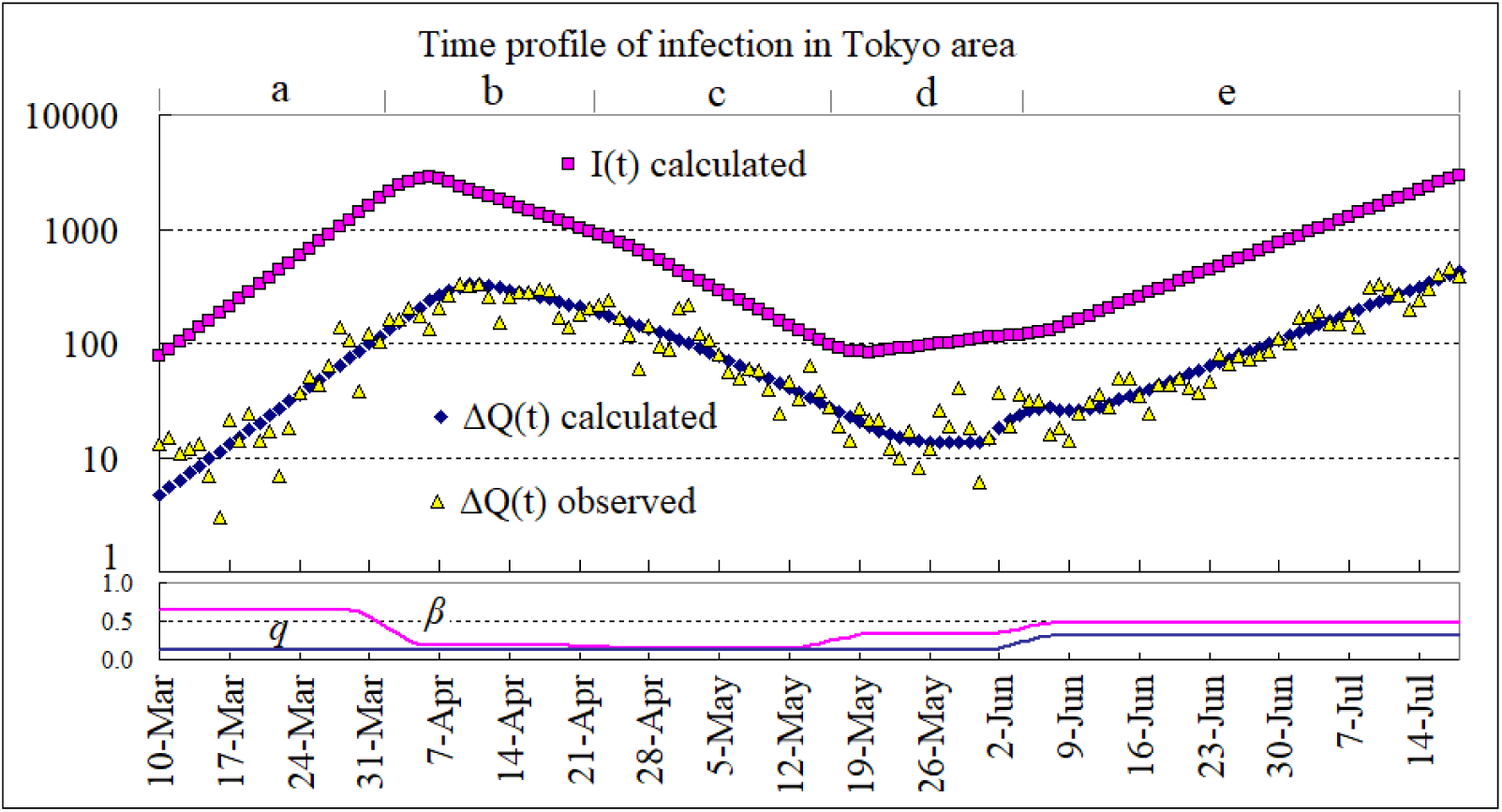
Time profile of infection in Tokyo area. Vertical axis is a common logarithm. The observed data are from public data [18]. *I*(*t*) denotes the number of infectious patients not quarantined. Δ*Q*(*t*) denotes the number of daily PCR test positive patients. Symbols on the top represent five periods. Parameters *β* and *q* are shown underneath the main graph. The parameter *τ* is set to be 4 days.

**Figure 7.**
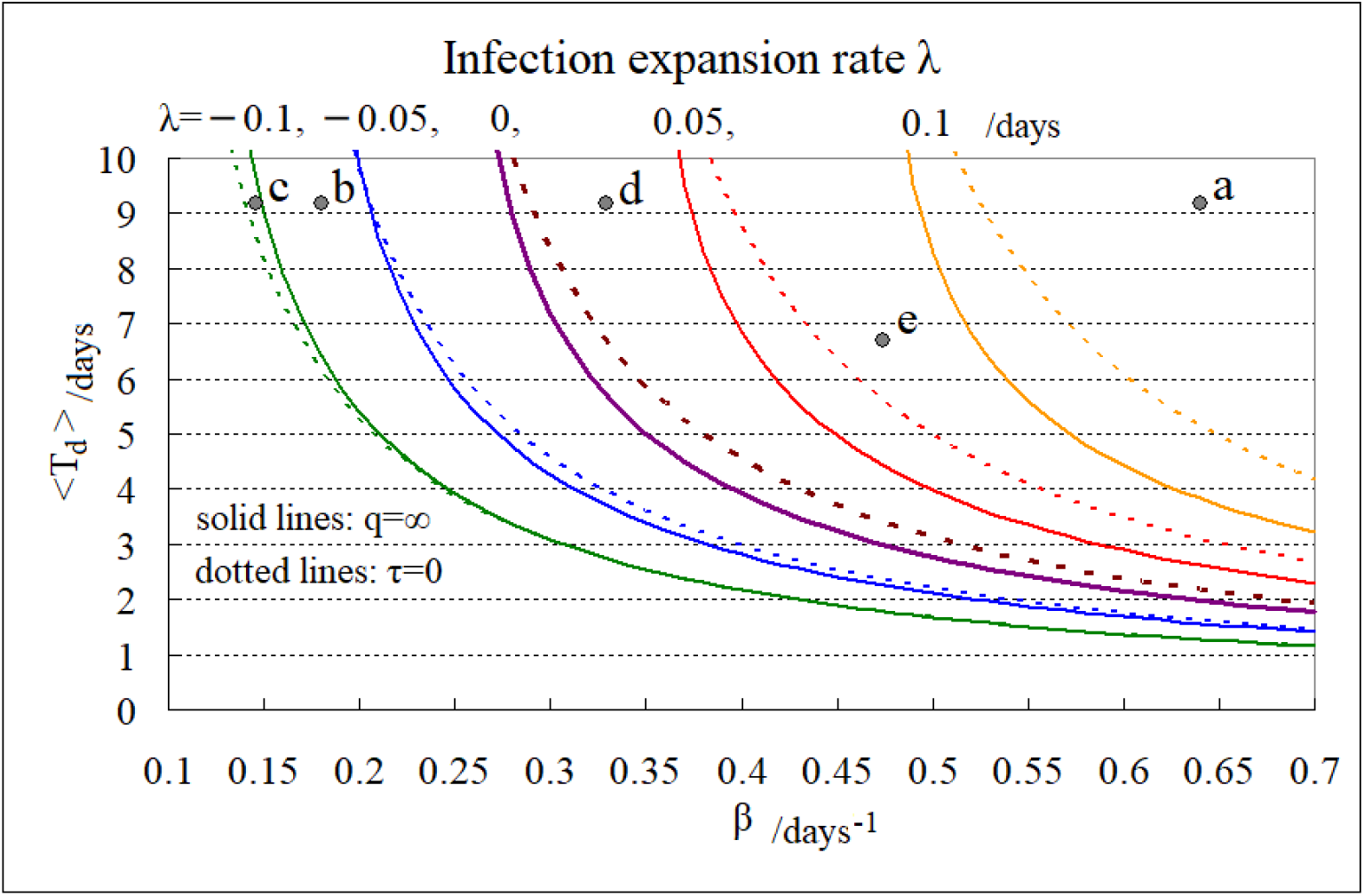
Infection expansion rate. The curves which make infection expansion rate *λ* =−0.1, −0.05, 0, 0.05, 0.1 are depicted in two cases. One is the case that *q*=∞ (*σ*=0), the other is the case that *τ*=0 (<*T*_d_>=*σ*).

This report has the following limitations.

1. I did not distinguish asymptomatic patients from presymptomatic patients. The data of asymptomatic case are very limited [22],[23],[24]. If both of its proportion and infectiousness are not negligible, it is necessary to take into account the difficulty of checking the asymptomatic case. One of the methods was developed in Appendix A, where some portion of patients can be left unchecked.
2. I assumed that the time profile of the onset of transmission (infectiousness) is an exponential type distribution. Actually it is estimated that there is a peak around the symptom onset time [3],[4]. This might affect the result when the average of quarantine delay is as short as the period of presymptomatic infectionon (2 days).
3. The symptom onset date was recorded only in 24% of the data. This loss of data might have an influence on the result of the estimation of the quarantine effect. Especially, I am afraid that there may be a tendency of forgetting to record the symptom onset date in the case of presymptomatic patient found by contact tracing. There would be a possibility of underestimate of the quarantine effect if the quarantine measure is further strengthened in future. A correct record on a symptom onset date is important to evaluate the quarantine effect.

## Data Availability

All the data that I used in the manuscript are available from the following link. https://gis.jag-japan.com/covid19jp/

## Acknowledgments

I would like to thank Drs. T. Odagaki and H. Mitsuishi for valuable discussion, and thank J.A.G Japan corporation for the epidemic data.

## Appendix A

Exact evaluation of the quarantine effect and the effective reproduction number

In order to reproduce exactly the same quarantine delay distribution as the observed one, we have to construct a different epidemic model similar to Young’s model [15]. The compartment of infectious patient I and that of the PCR test positive patient I’ are divided into many compartments J_τ_ and J’_τ_, respectively. The suffix τ means that all the members in that compartment are quarantined when *τ* days passed (corresponding to the case that *σ*=0 in the main text). (I do not need the compartment K nor K’.) In this report, I discarded the difficulty of checking asymptomatic patients. If we take that difficulty into account, we have to include a compartment with τ=∞. Equations (2.3),(2.4),(2,7) and (2.9) are replaced by the following equations.

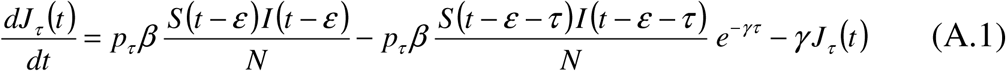

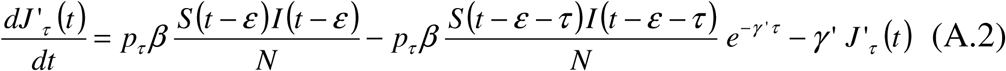

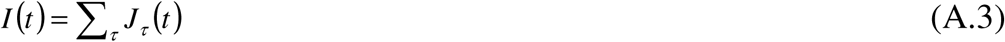

where *J*_τ_ denotes the number of the infectious patients and *J’*_τ_ denotes that of the PCR test positive patients in the compartments J_τ_ and J’_τ_, respectively. The parameter *p*_τ_ means the ratio of patients entering the compartment J_τ_ and J’_τ_. With an approximation that *S*=*N*, if a patient enters the compartment J_τ_ and J’_τ_ at *t*=0, the probability of being infectious without being quarantined is exp(−*γt*), and that of being PCR test positive is exp(−*γ’t*). Thus, the effective reproduction number is

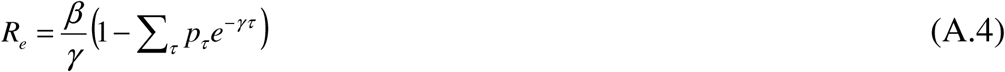

The time distribution of the number of quarantined PCR test positive patients is discrete.

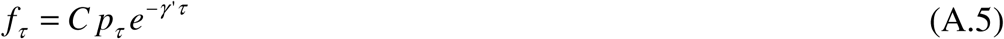

*C* is a normalization constant. Since there is no observed data corresponding to *f*_∞_, *C* should be determined as follows.

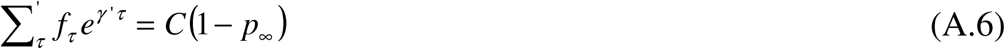

where Σ’_τ_ means sum with respect of *τ* <∞.

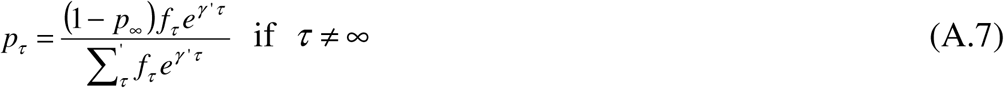

The effective reproduction number is expressed in terms of the time distribution of the delay of quarantine.

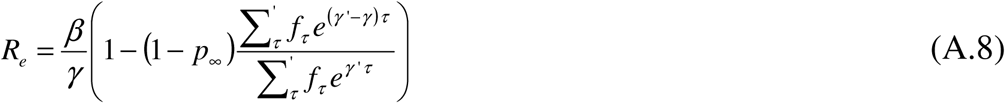

Equation (2.22) is obtained by assuming that *p*_∞_=0.

When the expansion coefficient *λ* is estimated from the time profile of the number of daily PCR test positive persons, the transmission coefficient *β* can be calculated from the following equation corresponding to Eq. (2.23).

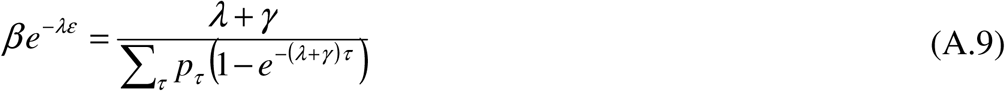

Therefore, knowing the time profile of the number of daily PCR test positive persons and that of the delay of quarantine, and neglecting the influence of asymptomatic infectious patients (*p*_∞_=0), we can derive directly the effective reproduction number using Eqs. (A.7) and the following equation.

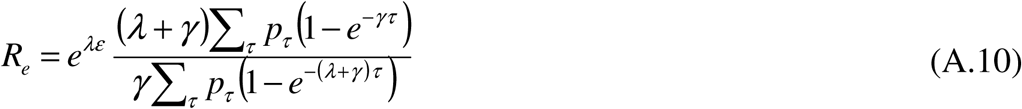

